# Counterproductive effects of Daratumumab and checkpoint inhibitor for the treatment of patients with relapsing NK/T lymphoma

**DOI:** 10.1101/2021.03.11.21251187

**Authors:** Wendy W.L. Lee, Jing Quan Lim, Tiffany P.L Tang, Daryl Tan, Puan Kia Joo, Kim Peng Tan, Liang Wei Wang, Ser Mei Koh, Choon Kiat Ong, Olaf Rotzschke

## Abstract

Natural killer/T cell lymphoma (NK/T L) is an aggressive malignancy associated with poor prognosis in relapsed patients. Although L-asparaginase based treatments are recommended as first-line treatment in relapsed patients, advances in immunotherapies such as checkpoint inhibitions have provided new therapeutic alternatives. However, as clinical outcomes for checkpoint inhibitors seemed to vary between NK/T L patients, combination therapies have been suggested to improve treatment efficacy. Here, we compared the effects of Daratumumab (anti-CD38)/anti-PD-1 combination therapy versus anti-PD-1 monotherapy on two relapsed NK/T L patients. Anti-PD-1 triggered an upregulation of CD38 on activated T cells, leading to depletion by Daratumumab. Concomittantly, EBV-specific antibody titer was also reduced alongside with depletion of CD38+ B cells and antibody-producing plasmablasts. Taken together, combining anti-CD38 and anti-PD-1 is likely to be antithetic.

## INTRODUCTION

NK/T-cell lymphoma (NK/T L) is a rare and aggressive form of an Epstein Barr Virus (EBV)-associated cancer with a predilection for non-Caucasian populations.[1] Most early stage patients were administered radiotherapy, which gives a 50% 5-year survival rate.[2] L-asparaginase based regimens such as SMILE (steroid, methotrexate, ifosfamide, L-asparaginase and etoposide) is given at late stages or relapsed patients. However, these chemotherapy regimes typically gave rise to adverse events such as high grade lymphopenia and increased infection risks, leaving SMILE refractory NK/T L patients with a poor prognosis.[3]

In recent years, advances in immunotherapy provide a new avenue for the treatment of cancer patients. By harnessing the body’s natural defence to fight cancer cells, this approach has proved to be effective for the treatment of many cancers including various forms of EBV-associated lymphomas. In particular, efficacy of checkpoint inhibition targeting receptor ligand pairs such as PD-1/PD-L1 monotherapy was demonstrated in patients with relapsed/refractory NK/T L, where ∼30-70% of the patients achieved complete response (CR) from PD-1 blockade therapy.[4-7]

PD-1 is a immunoinhibitory molecule expressed on activated lymphocytes including CD8+ T cells. Its partner ligand PDL1 is commonly found on antigen-presenting cells and lymphoma cells. Ligation of PD-1 with PDL1 inhibits T cell cytotoxicity [8, 9] and the use of anti-PD-1 such as Pembrolizumab perturbs this receptor/ligand interaction, hence unleashing the cytotoxicity of T cells to kill the tumour cells.[10]

Another way to obliterate cancer cells through immunotherapy is based on antibody-dependent cellular cytotoxicity (ADCC).[11] ADCC results in the depletion of cells that are bound by the targeting antibodies. For instance, anti-CD38 was found to effectively deplete multiple myeloma (MM) cells as these cells express high levels of CD38.[12] In another study, NK/T L cells were found to express CD38 and Daratumumab alone was highly efficacious in a patient with NK/T L.[13] Interestingly, CD38 is an activation marker that can be found on B cells, plasmablast and activated T cells.

In spite of the apparent effectivenes of immune checkpoint inhibition, it often works only in a subset of patients.[4-7] A common strategy to increase treatment efficiency is through the use of combination therapy to enhance synergistic effects.[14] However, not all treatments are compatible. Anti-PD1 and anti-CD38 monotherapy may have been effective for NK/T L patients. However, the effects of combining both treatment seems counterproductive as anti-CD38 treatment effectively eliminates key immune cell populations specifically activated by checkpoint inhibition. In this case study we demonstrate the neutralizing effect of Daratumumab on checkpoint inhibition by comparing immune cell composition, EBV DNA titer and EBV-specific antibody titers in blood samples of two relapsed NK/T L patients.

## METHODS

### Sample collection

Fresh blood samples were obtained from healthy volunteers and patients in accordance to the Helsinki declaration and approved by the SingHealth Centralised Institutional Review Board CIRB Ref: 2017/2806 and 2004/407/F respectively. Written informed consent was obtained from volunteer donors prior to sample collection.

### Quantitation of EBV genomic DNA levels in patient plasma samples

Patient plasma was split into two fractions and treated either Buffer RDD (QIAGEN) or DNAse I (QIAGEN). DNA from the DNAse I-treated fraction represents only virion-encapsidated DNA while DNA mock-treated by Buffer RDD represents total DNA from a patient’s plasma (cell-free host DNA, virion-encapsidated DNA and free-floating viral DNA). An equal volume of cold phenol-saturated Tris-EDTA (10 mM Tris, pH 8.0 with 1 mM EDTA) was added to each tube prior to vigorous shaking. Samples were then centrifuged at maximum speed at 4°C for 15 min. An additional 100 µL of Buffer EB (QIAGEN) was gently added to facilitate aspiration. The aqueous phase of DNA was extracted and stored at −20°C until use. Standard ethanol precipitation using glycogen as carrier was performed to concentrate the DNA. Buffer EB (QIAGEN) was used to resuspend the purified DNA. Virus quantification was performed in accordance to published qPCR protocol using 1 µL template DNA.[15] Standard curve was generated using DNA from the Namalwa EBV-positive Burkitt’s lymphoma cell line.[16]

### Anti-EBV VCA IgG and total IgG quantification

Quantification of total IgG was performed according to manufacturer’s protocol using Human IgG ELISA quantification set (Bethyl Laboratories) with heat inactivated human plasma samples diluted at 1:50,000. Quantification of anti-EBV VCA IgG was performed according to manufacturer’s protocol using EBV-VCA IgG ELISA kit (Calbiotech) using heat inactivated human plasma samples.

### Immunophenotyping by mass cytometry

PBMCs were isolated using Ficoll-Paque density gradient centrifugation, frozen in freezing medium (10% DMSO, 90% fetal bovine serum(FBS)) and stored at −80°C until use. For Immunophenotyping by mass cytomety, PBMCs were thawed using complete RPMI (RPMI+10%FBS) and washed with PBS. The cells are then treated with cisplatin for 5 mins, followed by incubation with metal-conjugated surface antibodies cocktail containing CD45, CD19, CD20, CD27, CD38, CD3, CD45RA, CD4, CD8 for 30mins on ice. Cells were washed twice with CyFACS buffer (PBS with 4% FBS, 0.05% sodium azide) and fixed overnight using 2% PFA made in PBS. The next day, cells were barcoded and stained with Cell-ID Intercalator-Ir (Fluidigm) in PBS for 20 mins at room temperature. Cells were washed twice with CyFACS buffer followed by a final wash using MiliQ water and passed through size filter. Filtered cells were acquired using Helios mass cytometer (Fluidigm) on CyTOF software version 7.0.5189. Analyses were performed using Flowjo V10.5.3

## RESULTS

### Treatment regimes

For this study we longitudinally tracked two relapsed NK/T L patients who failed SMILE therapy (Figure 1A). The first patient underwent anti-PD1 monotherapy (pt_MT) received Pembrolizumab on 21 and 24 days post SMILE treatment (dps) while the other patient who underwent combination therapy (pt_CT) was treated on day 42 dps with Daratumumab followed by subsequent treatment with anti PD-1 (Nivolumab) on 91, 105 and 120 dps. Pt_CT also developed transient leukocytosis from 117-128 dps, before demise at 176 dps. For this study, blood samples were obtained at days 21 and 24 dps for pt_MT and on 91 and 105 dps for pt_CT.

**Figure 1.**
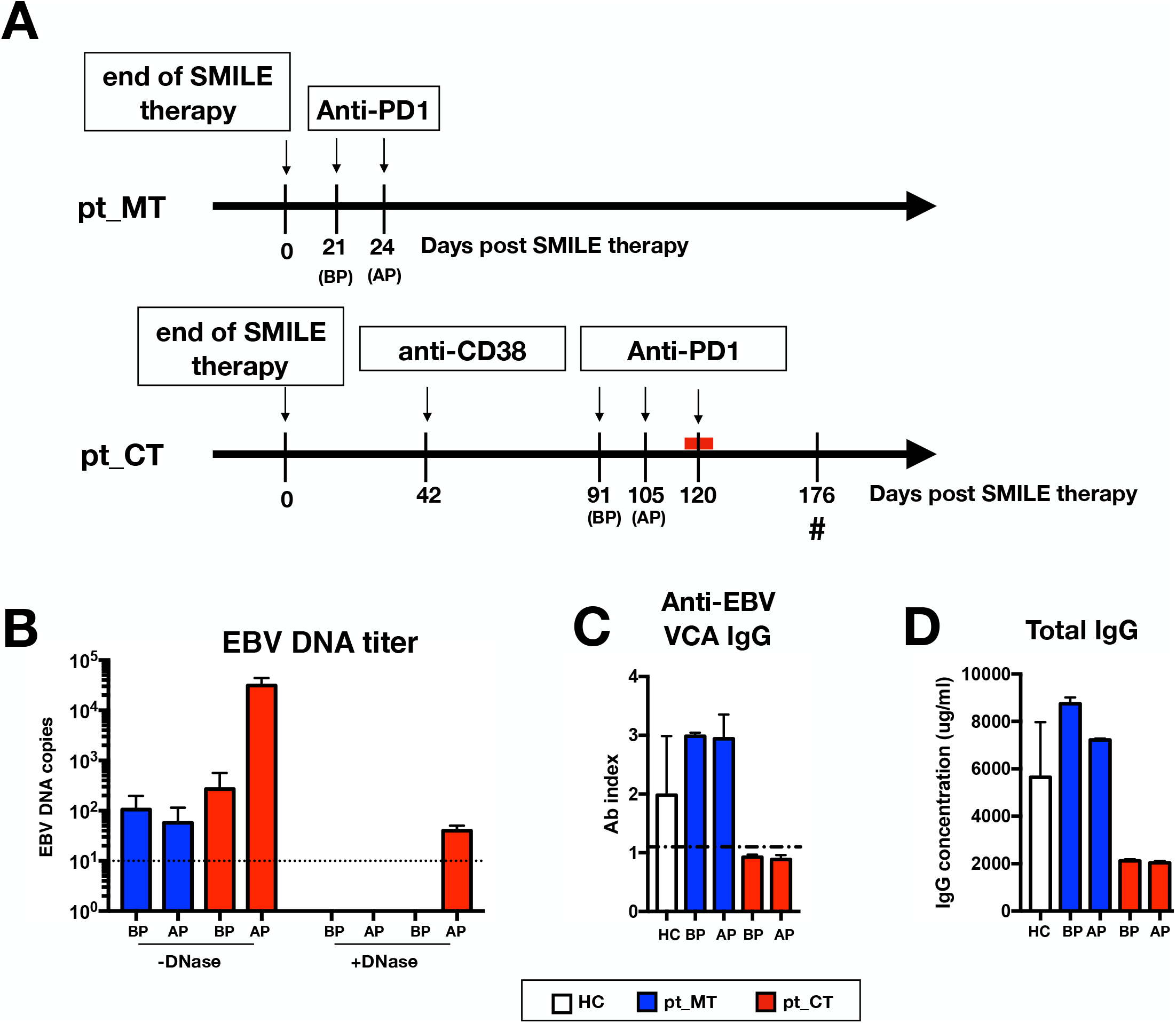
Combination therapy reduces EBV-specific and total immunoglobulin. (**A**) Timeline of treatment in patient undergoing anti-PD1 only monotherapy (pt_MT) and patient undergoing anti-CD38 followed by anti-PD1 combination therapy (pt_CT). pt_CT developed transient leukocytosis (indicated by red line on timeline) during the course of anti-PD1 treatment. BP and AP indicate timepoint before and after anti-PD1 treatment respectively. Hash (**#**) indicates demise of patient pt_CT. Numbers below the timeline shows the number of days after SMILE therapy was stopped for the respective patient. (B) Plasma samples were obtained from healthy donors (HC), pt_MT and pt_CT, at timepoints before (BP) and after anti-PD1 (AP) treatment. Bar charts showing **(B)** EBV viremia (with and without DNase added during sample extraction) and **(C)** levels of anti-EBV VCA IgG. **(D)** Total IgG quantification was also performed in HC and patients.

### Increased EBV viremia and reduced levels of EBV-specific IgG in patient undergoing combination therapy

As NK/T L cells release EBV DNA, the tumour burden can be directly associated with the amount of EBV DNA circulating in the blood.[17] At the start of the Pembrolizumab treatment, comparable levels of serum EBV DNA detected in both patients (Figure 1B). While in patient pt_MT, a slight reduction in the level of circulating EBV DNA was observed after anti-PD1 treatment, a marked increase was detected in patient pt_CT (Figure 1B). Further analysis shows that a small percentage of the viral DNA in pt_CT is derived from virions (DNase-protected), suggesting that the increased level of circulating DNA are likely at least in part due to reactivated virus replication. Notably, virion-encapsidated EBV DNA was not detected in the case of pt_MT.

It is estimated that 90% of the world population has been infected by EBV.[18] Thus, it is expected that most of the healthy controls have high levels IgG against EBV (Figure 1C). Equally high levels of anti-EBV IgG were also found in patient pt_MT (Figure 1C). Surprisingly, patient pt_CT was found to be negative for anti-EBV IgG (Figure 1C). Notably, while EBV-specific IgG was undetectable, the patient was also found to have low levels of total IgG (Figure 1D).

### Depletion of antibody producing plasmablasts by anti-CD38

The low levels of EBV-specific and total IgG in patient pt_CT (Figure 1C and D) motivated us to check whether B cells are affected in this patient. Peripheral blood mononuclear cells (PBMCs) from the two patients were analysed using flow cytometry. Healthy donors were included as controls. Prior to anti-PD1 treatment, both patients had substantially reduced B cell numbers as compared to healthy controls (Figure 2A). However, patient pt_MT showed a marked increase in CD38+CD27+ B cells, which consist of plasma cells, antibody-producing plasmablasts and memory B cells (Figure 2B and C). Patient pt_CT in turn showed a striking depletion of all CD38+ B cells to negligible levels. This was apparent in both CD38+CD27+ (Figure 2B and C) and CD38+CD27-(naïve and transitional) (Figure 2B and D) B cell populations. The analysis for pt_CT was done nearly 50 days after the anti-CD38 injection, stressing the efficiency of the cell depletion by Daratumumab. This was in line with another study which shows the effects of Daratumumab can persist up to 3 months post administration [19].

**Figure 2.**
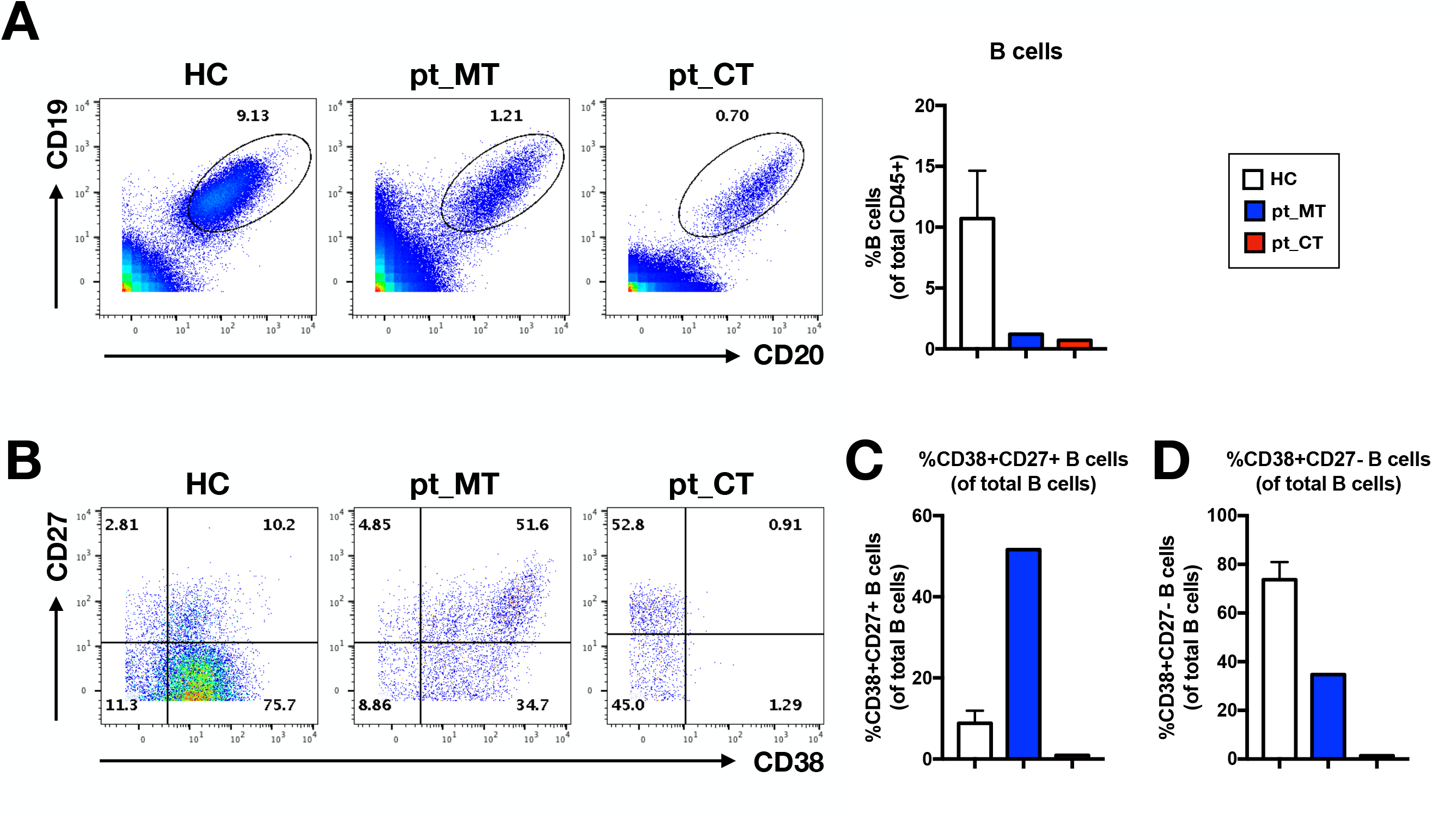
Combination therapy depletes CD38+ B cells subsets. PBMC samples were obtained from HC, pt_MT and pt_CT, at timepoints before (BP) and after (AP) anti-PD1 treatment. Immunophenotyping of the PBMCs was performed. **(A)** Scatterplots (left) and barchart (right) showing percentage CD20+CD19+ B cells of total CD45+ leukocytes. Number in the scatterplot show the percentage of CD20+CD19+ B cells of total CD45+ leukocytes. **(B)** Scatterplots showing CD27 and CD38-expressing B cell subsets. Numbers in the scatterplot show the percentage of respective population within each quadrant of total B cells. Bar charts showing the percentage of **(C)** CD38+CD27+ B cells and **(D)** CD38+CD27-B cells of total B cells derived from **(B)**.

### Anti-CD38 therapy depletes the T cells activated by anti-PD1

Given the effective depletion of Daratumumab on CD38+ antibody producing B cells, it is likely that it will have a similar effect on the T cell compartment. One of the desired outcomes for anti-PD1 treatment is to reinvigorate exhausted tumour-specific CD8+ T cells so that they can directly kill tumour cells. Comparative analysis of samples from patient pt_MT taken before and after anti-PD-1 treatment confirmed the expansion of CD45RA-T cells (Figure 3A). Moreover, the analysis further revealed that these CD45RA-cells also strongly upregulated CD38.

**Figure 3.**
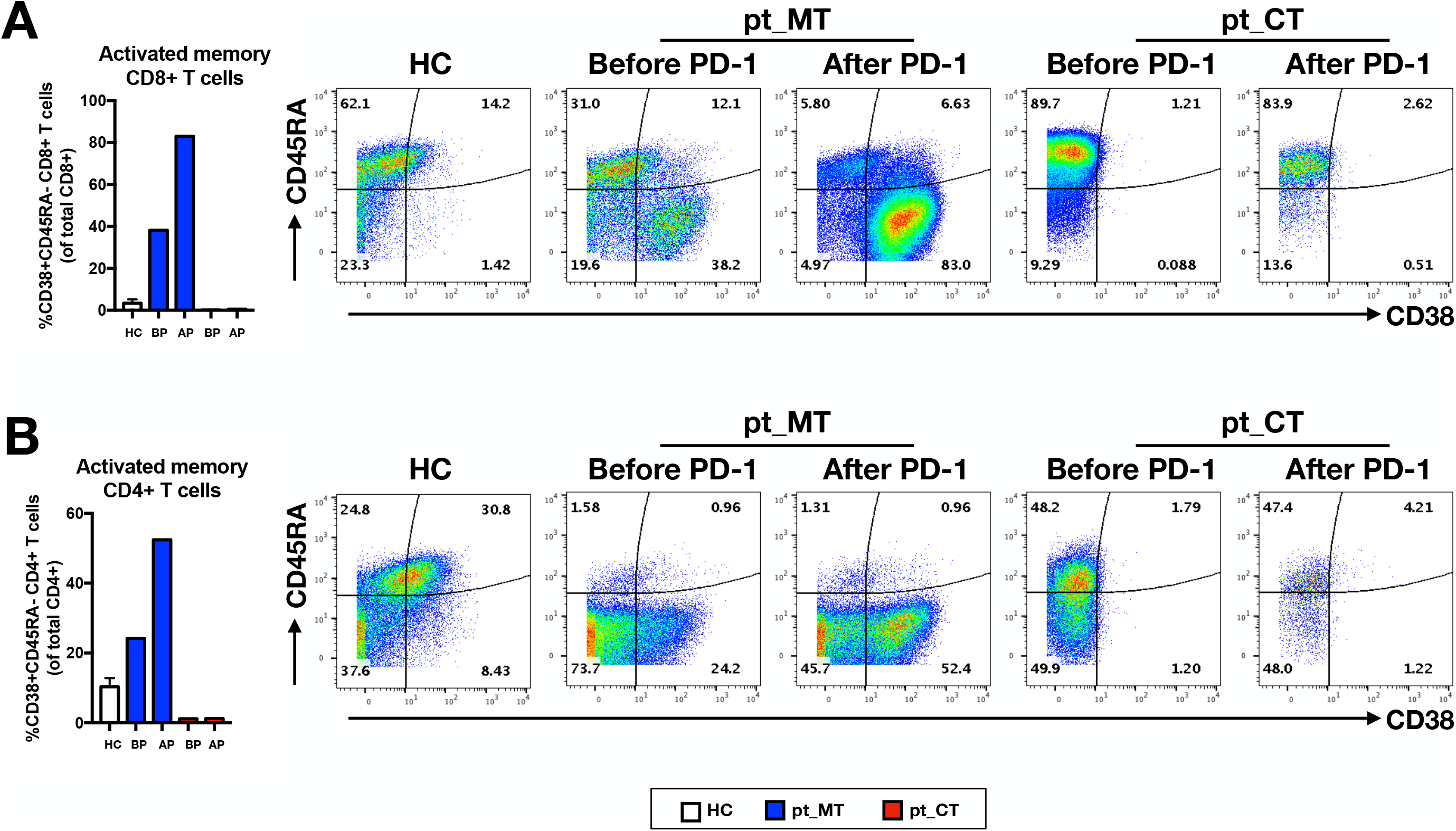
Combination therapy abate activated CD38+ T cells. PBMC samples were obtained from HC, pt_MT and pt_CT, at timepoints before (BP) and after (AP) anti-PD1 treatment. Immunophenotyping of the PBMCs was performed and T cells populations were analysed. **(A)** Bar chart (left) showing the percentage of activated memory CD38+CD45RA-CD8+ T cells of total CD3+ T cell population. Scatterplots (right) show CD45RA and CD38-expressing CD8+ T cells. Numbers in the scatterplot show the percentage of respective population within each quadrant of total CD8+ T cells. **(B)** Bar chart (left) showing the percentage of activated memory CD38+CD45RA-CD4+ T cells of total CD3+ T cell population. Scatterplots (right) show CD45RA and CD38-expressing CD4+ T cells. Numbers in the scatterplot show the percentage of respective population within each quadrant of total CD4+ T cells.

Prior to the checkpoint inhibition, about 38% of the CD8 T cells were of CD45RA-CD38+ activated phenotype and this proportion further increase to more than 80% following anti-PD1 treatment (Figure 3A). On the other hand, a striking absence of CD38+ CD8+ T cell was observed in patient pt_CT (Figure 3A). This lack of CD38+ T cells were concordant before and after anti-PD1 treatment, suggesting that even 50 days after injection there are still sufficient amounts of the anti-CD38 antibody in the circulation to deplete all cells expressing the surface marker as a result of the checkpoint inhibition. Intriguingly, this observation was also made in the CD4+ T cells compartment (Figure 3B).

## DISCUSSION

Checkpoint inhibition therapy such as anti-PD1 and anti-CD38 depletion therapy have both been suggested to be alternative treatments for late-stage NK/T L patients that are refractory to chemotherapy.[4-7, 13] Checkpoint inhibition therapies works by preventing inhibitory receptor signals of tumour-specific immune cells to revigorate their cytotoxic capacity and has been shown to be effective for the treatment of tumours with high neo-antigen load.[20]

Recent studies have shown that anti-PD1/PD-L1 treatment is also effective for NK/T L.[4-7] In line with these reports, we observed that patient pt_MT responded well to anti-PD1 treatment in terms of clinical progression as well as changes in the PBMC composition. Administration of anti-PD1 triggered a massive activation and expansion of CD45RA-effector/memory cells on both cytotoxic CD8+ as well as helper CD4+ subsets in the patient.

On the other hand, anti-CD38 depletion which directly target the tumour via ADCC was also shown to be an alternative treatment for NK/T L as high expression of CD38 was found on the tumour cells. Daratumumab was reported to have some efficacy in depletion of the lymphoma.[13]

While it has been suggested that a combination of these two approaches could further enhance treatment efficacy,[21] here we present data from two cases of NK/T L relapsed patients who have undergone anti-PD1 treatment and one of them with anti-CD38 treatment prior to anti-PD1 treatment, to show that the combined use likely to be counterproductive.

In addition to NK/T L, CD38 is also an activation marker expressed on activated T cells in response to anti-PD1 treatment. The importance of these CD38+ leukocytes was observed in hepatocarcinoma patients, where patients with higher levels of infiltrating CD38+ cells were found to respond better to anti-PD1 treatment.[22]

By monitoring the blood profile of patient pt_CT, we demonstrated that anti-CD38 treatment adminstered prior to anti-PD1 treatment depleted key immune populations activated by anti-PD1 treatment. Activation of T cells by anti-PD1 resulted in the upregulation of CD38 and predisposes these cells to depletion by the circulating Daratumumab antibodies, thus abrogating any beneficial effect that may have arisen from the checkpoint inhibition. Although the case of death may be unrelated to the treatment, patient pt_CT succumbed to the disease shortly.

Daratumumab has shown clinical efficacy in the treatment for multiple myeloma (MM) as these cancer cells expresses high levels of CD38.[12] MM arises from plasma cells, which are antibody-secreting B cells which generally express high levels of CD38.[23] It is thus unsurprising to find patient pt_CT to be severely depleted of

CD38+ B cells, which include plasma cells and plasmablasts. Depletion of these cells also accounted for very low total IgG levels and in particular, for the absence of anti-EBV IgG titer despite the high levels of circulating EBV DNA in the patient. The small burst of virion production as evidenced by the detection of encapsidated viral DNA may be detrimental as there is potential for secondary EBV infection of residual B cells. The loss of antibody titers and B cell responses can also cause patients to be more susceptible to infections, which could be a contributing factor to the uncontrollable increase in EBV viremia in patient pt_CT. However, this observation contradicts the results from an MM study, where no reduction in IgG levels were seen in patients treated with Daratumumab.[24] It thus remains to be seen to what extent Daratumumab compromises the humoral response.

Despite the small sample size, this study has important clinical implications in the treatment of NK/T L patients as it provided strong evidences to caution against the combined use of anti-PD1 and anti-CD38 agents, despite their successes in single-agent clinical studies.

## Data Availability

All data in the manuscript are available

## ACKNOWLEDGEMENTS

We thank all patients and healthy donors for making this study possible.

